# Using epidemic simulators for monitoring an ongoing epidemic

**DOI:** 10.1101/2020.05.08.20095463

**Authors:** Mohan Raghavan, Kousik Sarathy Sridharan, M R Yashaswini

**Affiliations:** Department of Biomedical engineering, Indian Institute of Technology, Hyderabad, Hyderabad, India

**Keywords:** COVID-19, SARS-CoV-2, contact tracing, testing, epidemiology, compartment models

## Abstract

Prediction of infection trends, estimating the efficacy of contact tracing, testing or impact of influx of infected are of vital importance for administration during an ongoing epidemic. Most effective methods currently are empirical in nature and their relation to parameters of interest to administrators are not evident. We thus propose a modified SEIRD model that is capable of modeling effect of interventions and in migrations on the progress of an epidemic. The tunable parameters of this model bear relevance to monitoring of an epidemic. This model was used to show that some of the commonly seen features of cumulative infections in real data can be explained by piece wise constant changes in interventions and population influx. We also show that the data of cumulative infections from twelve Indian states between mid March and mid April 2020 can be generated from the model by applying interventions according to a set of heuristic rules. Prediction for the next ten days based on this model, reproduced real data very well. In addition, our model also reproduced the time series of recoveries and deaths. Our work constitutes an important first step towards an effective dashboard for the monitoring of epidemic by the administration, especially in an Indian context.

## Introduction

Mathematical treatment of epidemics has its origins more than a century ago [1, 2]. The recent interest in modeling epidemics was rekindled with the onset of the Severe Acute Respiratory Syndrome (SARS) epidemics during the early part of the century [3]. The classical model of epidemics (SIR) was designed as a dynamical system with the fraction of Susceptible(S), Infected(I) and Recovered(R) population as state variables. The SARS epidemics were modeled with a modified model with an additional state for the fraction of exposed population(E) that is latent and as yet uninfected, but still contributes to spreading the disease. This modified model, the SEIR model, is currently being used to characterize the epidemics caused by a group of coronaviruses including the ongoing COVID-19 [3]. Modified SEIR models have also been designed to understand the effect of quarantining [3] and multiple active strains of the pathogen simultaneously active in populations [4]. SIR, SEIR and related models are also known as compartment models, as population groups are modeled as a single compartment. In contrast, agent-based approaches, model the transitions of individual agents in a population and hence can capture the effects of non-homogenous populations. While agent based and stochastic models [5] have been explored, it has been shown that for people to people contact networks that are small world, random, fully connected or scale free, the compartmental models work quite well [6]. Thus, compartmental models are an efficient tool for modeling epidemics.

Some of the important parameters that characterize an epidemic are the rate at which new infections are created (*β*), mean duration of infection (*T_inf_*) and their product (*R*_0_), which gives the mean number of infections spawned by an individual during the infectious period when the entire population is susceptible. The emergence of *R*_0_ in epidemiology owes its origins to the mathematics of population growth, and in past few decades, emerged as an important diagnostic of the epidemic progression [7, 8]. *R*_0_ owes its importance [9] to the fact that, it is a prognosticator of the epidemic, with values less than 1 indicating a fade out and values greater than 1 prognosticating a large scale epidemics. *R*_0_ can easily be calculated from the set of differential equations describing SIR and SEIR systems [7].

The method can be applied with modifications for calculating the reproduction numbers for extended models with more number of compartments if they satisfy certain conditions[10]. While *R*_0_ is the value of *R* at the start of the epidemic, *R_t_* represents its evolution with time *t*. The change in *R_t_* with time could be due to a number of factors, namely a change in transmission *β* or an intervention in the form of lockdowns, social distancing, testing and / or quarantining. While the dynamics of transmission of epidemic diseases are well understood, estimating these parameters during the course of an ongoing epidemic is not trivial, except in special circumstances of small isolated population clusters as in the quarantined cruise ship [11]. For the purposes of monitoring and predicting the course of an ongoing epidemic, the most important metric used is *r*, the slope of the cumulative infections on the log scale. This follows from the observation that the rate of new infections is proportional to the cumulative infections. The parameters *r* and *R* are related to each other through the shape of the serial interval distribution [12, 13]. Since the cumulative deaths follow the infections with a lag, the cumulative death count has also been used to estimate the parameters of the epidemic model [14]. *R* has also been estimated using maximum likelihood based methods from the serial intervals distribution for subsets of populations where the chain of infection transmission are documented [15].

Methods used to recreate the time varying parameters underlying an epidemic, in particular COVID-19 are largely of two kinds. First, the class of mechanistic models which use modified SEIR compartment models to simulate epidemics. The second, is empirical in nature and rely on fitting of parameters of mathematical formulations to real data. The mechanistic models are largely used to evolve strategies for management of the epidemic - like evaluate effect of social distancing, lockdowns, exits from lockdowns [16], projection for periodic recurrence of outbreaks and impact of multiple strains of the pathogen [4]. The parameters and formulations used in these methods are grounded in epidemiology theory and are useful to evolve broad strategies. But they are not frequently used for short or medium term prediction of trends or evaluation of efficacy of interventions in real time. For such predictions, the second class of empirical methods are deployed. One particular study modeled the transmission as a geometric process. Using Monte Carlo simulations, *R_t_* and percentage of undetected infections were estimated by fitting to epidemiological data from Wuhan [17]. Another study used convolution of daily reported infections with the serial interval distribution to estimate the cumulative number of infections and deaths. Interventions are modeled as contributing factors to a country specific, time varying *R_t_*. The contribution of various interventions were estimated by fitting to data [14].

On the issue of estimating epidemic parameters certain salient problems limit efficacy of current methods. Foremost among these are the gap between observed and true variables like infections and recoveries. This is less of a problem in case of deaths, as they are not easily hidden, which was used in estimation [14]. However, it requires about 2-3 weeks for the effect of interventions to be reflected in the number of deaths, which severely limits the utility of the method for real time analysis. Empirical methods have a general limitation that they make assumptions on various parameters like serial interval distributions or the form of priors [14, 17]. Due to the lack of mechanistic details, the results of these methods are often incapable of offering intuitive explanations on how interventions work or fail. Mechanistic methods require that standard models be modified to incorporate interventions, testing efficacies and quarantining often making estimation of *R_t_* very difficult. Over and above all of these, we believe that a common shortcoming in most mechanistic models proposed are a conflation of observables and non-observables. For instance, the deaths and recoveries are almost always modeled as single compartments and outcomes resulting from both detected and undetected infections move into the same compartment, although one is observable and the other is only partially so [3, 4, 16, 18].

Thus, there is a clear need for models that can explain short and medium term trends in epidemic data while providing clear mechanistic link to the effect of interventions amidst epidemics. We propose a mechanistic model that retains the simplicity of the standard SEIR model, but with the power to model interventions and time varying transmission. A key constraint we impose on our model is that while it should be able to explain short term trends and effects of interventions, it must also be capable of providing intuitive connections between observed trends and changes in underlying parameters. In this work we present one such SEIR model suitably modified to model interventions. We then proceed to demonstrate how changes in model parameters can generate oft-seen patterns in cumulative infections on the logarithmic scale. Based on these results, we propose heuristic algorithm for estimating time varying interventions that recreate trends in cumulative infections by applying suitable interventions, thereby estimating the underlying changes in epidemic dynamics. We use this method to recreate the real data from 12 Indian states between mid-March and mid-April. We finally observe how this model fares in predicting the trends of the succeeding 10 day period.

## Methods

The key principles for the design of the epidemic simulator were as follows:

- **Less is more:** We choose to represent just enough compartments as required. A compartment or a transition that neither has a correlate in real data nor can be used to estimate an observable and useful quantity is discarded or merged.
- **Simplicity**: Model must use as little sophistication as possible. Expressions for key quantities such as reproduction number must remain as close as possible to the standard SEIR models.
- **Intuition:** An administrator should be able to intuitively identify the parameters used in the model with interventions deployed on the ground in tackling the epidemic.

### Model Design

#### Compartments

We use a modified version of the SEIRD model on the lines of [3]. We duplicate the standard SEIRD pipeline to create two parallel pipelines - undetected and detected / isolated. In particular, we also duplicate the deceased and recovered compartments in both pipelines.

The motivation for the same is as follows. Daily confirmation of cases, recoveries and deaths are both released and tracked. These numbers count only the detected infections and their recoveries. However number of fatalities can come from amongst the detected and undetected infections. Recently the Indian government has mandated COVID-19 screening for all cases with severe acute respiratory infections. These are with a high probability critical cases and many succumb. These numbers are often added to count of deceased, but in an inconsistent manner. Thus the trends of reported deaths would tend to fluctuate between true death numbers and those coming from the detected infections.

We do not use other possible sub compartments within the infected - like mild infection, hospitalized, critical etc. While we do believe that these are important, the number of hospitalizations and grades of infections are not available in the public domain. Separate compartments for quarantined susceptible are important when estimating the cost, and resources spent due to numbers of people that are quarantined although not exposed. Since we do not, at the moment consider this aspect, we dispense with a separate compartment for susceptible-quarantined.

Thus we arrive at a total of 9 compartments which include two compartments each for Exposed, Infected, Recovered and Dead and a single compartment for susceptible.

#### Interventions

We recognize two interventions commonly deployed for identifying infections as part of epidemic management.

- **Contact tracing and isolation:** When an infection is detected, immediately their contacts are traced and placed under quarantine. They are tested for infection periodically until they test positive or the observation period ends. Within our model, we represent the efficiency of this intervention by the fractional value *c*, which represents the fraction of exposures and transmitted infections that are detected and quarantined. This parameter lends itself to model simplicity, yet intuitive and actionable for administrators.
- **Random testing and self-presentation:** A patient voluntarily reports symptoms and is then tested. Alternately, random tests are administered to the public and positive reports are quarantined. Within our model, we represent the efficiency of this intervention by the fractional value *q*, which represents the fraction of infections that are detected and quarantined.

We assume that during the exposed phase, people are largely asymptomatic and undetectable by tests. Thus the exposed are quarantined only by contact tracing. However, the infected can be detected by either contact tracing or random tests / self-reporting. Further we make a simplifying assumption that during stage 2 of an epidemic, detection is mostly due to contact tracing and during stage 3, when community transmission takes over, detection of infections from random testing or self-reporting far outnumber the ones due to contact tracing.

#### Transitions and flow of people between compartments

In order to keep the model simple enough for numerical analysis, we employ two rules for designing transitions

- All transitions must follow the same order as S → E → I → R. For instance, all exposed compartments or their sub compartments (quarantined or undetected) must have transitions only to an infected compartment or one of its sub compartments.
- When there are multiple transitions out of a compartment, the sum of the rates out of the compartment must be conserved (compared to un-branched rates)

Starting from the standard SEIRD model, we observe that the exposed population, consists of 2 sub populations – the quarantined and free. Since quarantining of exposed is only due to contact tracing, a fraction c of the exposed flow to the compartment *E_q_*. Thus we split the *S → E* transition into two transitions *S → E_q_* and *S → E*. The rates of these transitions are in the ratio *c*:: (1 − *c*) and the sum of their rates continues to be *β.S.I* as in the standard SEIRD model. It must be noted that I in the standard model includes all infected, while here it refers only to the undetected infections. See Fig. 1 for more details.

**Figure 1.**
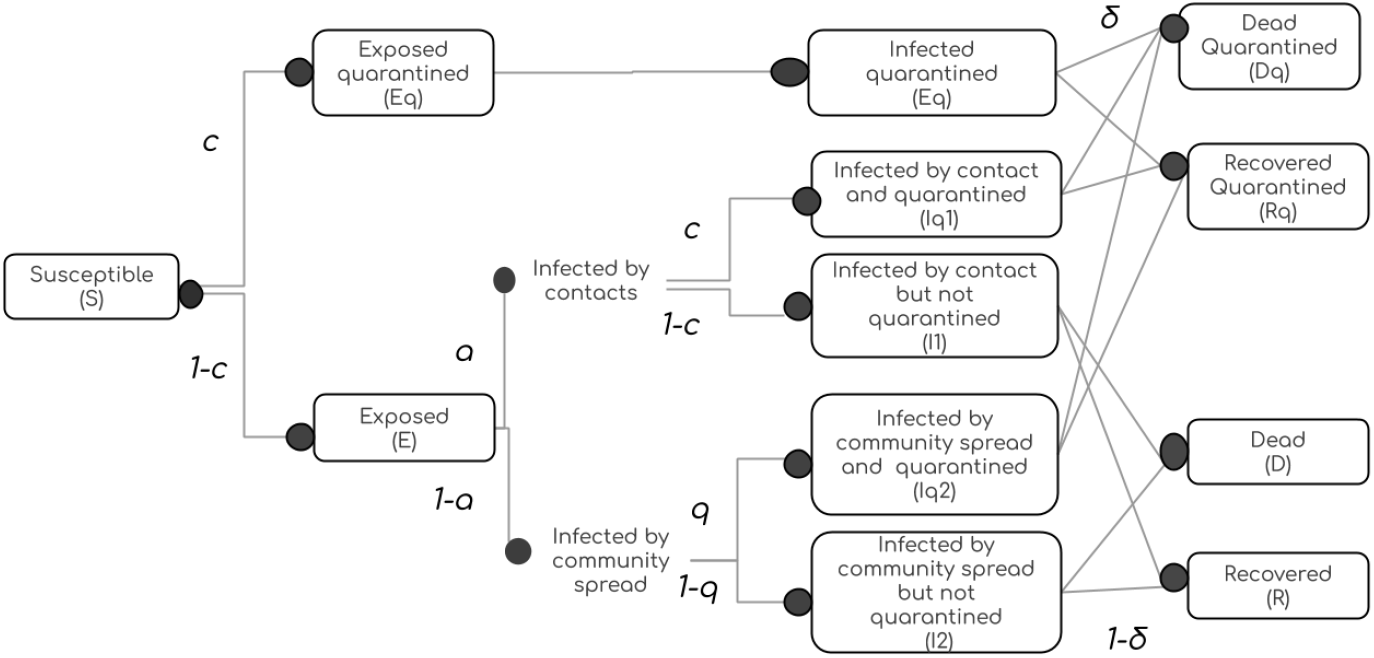
Flow of people through compartments. The compartments *E_q_, I_q_, R_q_, D_q_* represent the populations that are quarantined. Compartments *E*, *I*, *R* represent the exposed, undetected infections or recoveries. *D* represents the fatalities from the undetected population, but they may be detected close to death or post-mortem. *a* is the fraction of Infected that acquired it by direct contact with another infected. *c* is the fraction of exposed or contact infected individuals that are detected. *q* is the fraction of community infected that are detected by self reporting or random testing. *δ* is the rate of mortality.

Similarly, the infected population may be thought of as consisting of 2 sub populations – infected by direct contacts or through community spread. The fraction of contact spread infections and community spread infections are denoted by *a* and (1 − *a*) respectively. For simplicity we assume that only contact tracing is effective in detecting contact spread infections and only random testing or self reporting can detect community spread infections. This leads to 4 sub populations within the so far undetected infected population. Rates of transition to and fro the four infected populations *I_q_*_1_ through *I_q_*_4_ are computed as described before.

#### Migration and influx of infected

An important modification to standard models is necessitated by migrations. Influx of infected changes the seeding pattern of epidemic and plays a critical role in evolution of epidemic. Influx of infections are represented in our model as a combination of inflow rate and probability that an incomer is infected. Given the inflow rate is *ϕ* and the probability of infection is *pI*, in-migration is implemented as an addition of 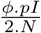 to the undetected *E* and *I* compartments each, per day. We assume that this inflow is finite and very small compared to the total population and hence the stable population assumption is not violated.

#### Model Parameters

It may be noted that because of the way transitions were designed in section on *Transitions*, the net rates of transition, Rate{*C*_1_ → *C*_2_} ∀ *C*_1_, *C*_2_ ∊ {*S*, *E, I, R, D*} remain the same as in the standard SEIRD model when the compartments are interpreted as unions of their sub compartments, i.e. *C* = ⋃ {*C, C_q_*}. The mean stay times in an Exposed (*E*) state and Infected (*I*) state were set to 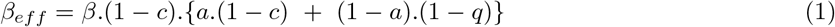 *days* and 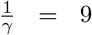 *days* respectively. The mortality rate *δ* is a constant value for the entire duration of simulation. Permissible values are in the range 5% ± 3%. The rate of transition of *S* → *I* is proportional to the fraction of total population that is infected (*I* + *Iq*) and the transmission rate (*β*). But since *I_q_* is quarantined and cannot influence this rate, only the undetected infections *I* contribute to this rate. Only a fraction (1 − *c*) of possible exposures reach *E*. Out of this number, a fraction *a*.(1 − *c*) are contact transmission cases and missed by the contact tracing exercise. A fraction (1 − *a*).(1 − *q*) of the undetected exposed are community transmitted infections and are missed by random testing. Thus the net effective rate of the transition *S* → *E* (union of *E* and *E_q_*) under the influence of interventions is given by *β*.*I* or *β_eff_ ·*(*I* + *I_q_*), where

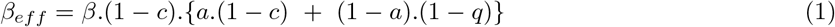

*β_eff_* is the effective beta in the standard model that corresponds to modified model with interventions. *β_eff_* is *β* scaled by the product of inefficiencies of two phases of interventions - isolation of exposed and isolation of the infected. The inefficiency of detecting the exposed is given by (1 − *c*) as testing cannot detect them and contact tracing is the only effective measure. The inefficiency of the detecting the infected is a convex combination of the inefficiencies of contact tracing (1 − *c*) and random testing or self reporting (1 − *q*). In stage 2 it is given by (1 − *c*) and in stage 3 by (1 − *q*).

The net transitions out of *E* and *E_q_* each continue to be *k*. The rates of *E → I_q_* are given by the sum of rates *E* → *I_q_*_1_ and *E* → *I_q_*_3_. The rates of *E* → *I* are given by the sum of rates *E* → *I_q_*_2_ and *E* → *I_q_*_4_. These details are evident from the Fig. 1 and Fig. 2. For the SEIR model with a stable bounded population, time varying reproduction rate *R_t_* is given by 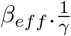. The term (1 − *c*).{*a*.(1 − *c*) + (1 − *a*).(1 − *q*)} that scales *β* and hence *R_t_* is the fraction of generated infections that go undetected. This fraction is an index of the inefficiency of interventions by the administration to control the epidemic. The inefficiency index assumes a simple form (1 − *c*)^2^ in stage 2 of the epidemic when contact transmission is the norm and takes the form (1 − *c*).(1 − *q*) during community transmission driven stage 3 of the epidemic.

**Figure 2.**
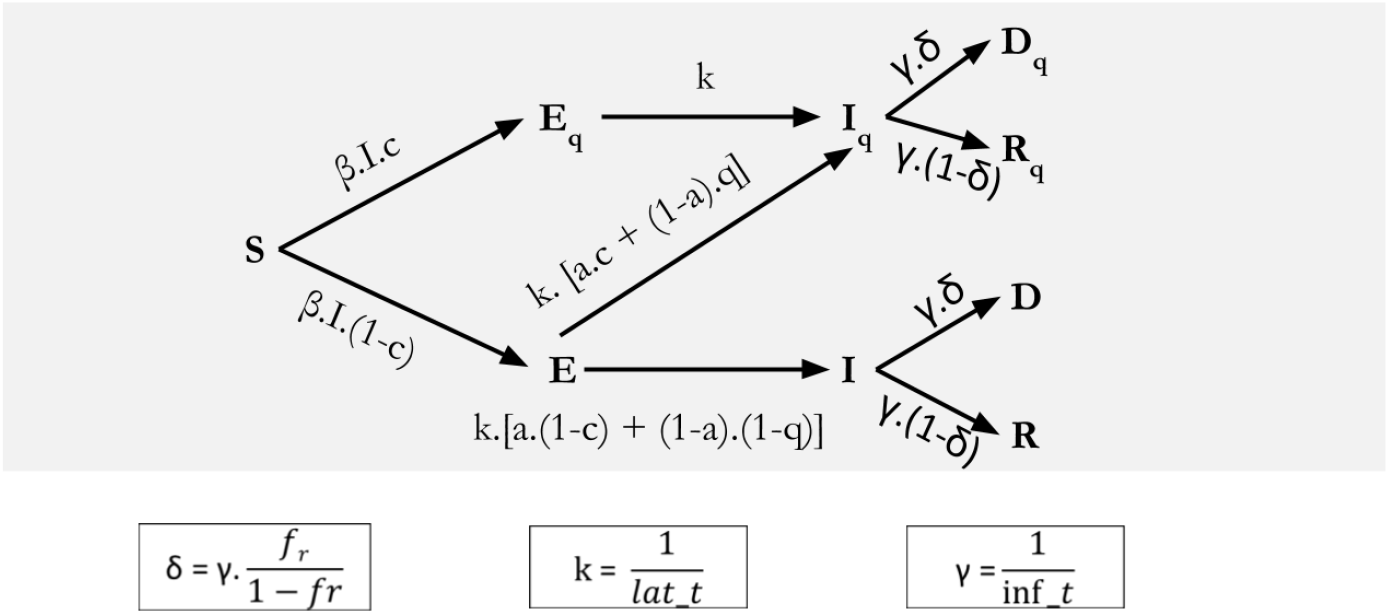
Modified SEIRD model. The compartments *E_q_*, *I_q_, R_q_*, *D_q_* represent the populations that are quarantined. Compartments *E*, *I*, *R* represent exposed, undetected infections or recoveries. *D* represents the fatalities from the undetected population, but they may be detected close to death or post-mortem. *a* is the fraction of Infected that acquired it by direct contact with another infected. *c* is the fraction of exposed or contact infected individuals that are detected. *q* is the fraction of community infected that are detected by self reporting or random testing. *δ* is the rate of mortality. the rates *γ* and *k* are respectively the inverses of mean infection time and mean latency times. *β* is the transmission rate given by product of contact rate and probability of infecting a contact

The values of *c*, *q*, *β* and *ϕ.pI* constitute the free variables of the models that are tuned to reproduce the true field data. Each of these quantities is assumed to be a piece wise constant function [14]. *δ* is a constant throughout a simulation and is chosen within the permissible range for each data set. *a* is of the form of a decreasing sigmoid function of the number of active infections. It has a transition from 0.99 to 0.01 between 0.2 * *e*^−4^ * *N* and 0.8 * *e^−^*^4^ * *N* numbers of actively infected.

#### Modelling response of Indian states

We use this model to estimate the dynamics of epidemic transmission underlying the time series of data from Indian states. Data of daily infections, recoveries and deaths between 13^th^ March 2020 and 17^th^ April 2020 for all Indian states were obtained from www.covid19india.org. We chose 12 states with active COVID-19 cases for our analysis. The states chosen were Delhi, Punjab, Uttar Pradesh, Rajasthan, Madhya Pradesh, Gujarat, Maharashtra, Telangana, Karnataka, Tamil Nadu, Kerala, and West Bengal. Using the data upto 17^th^ April 2020, we use a heuristic algorithm to generate these trends from our model. The algorithm will be derived from the results of model characterisation and will be described in section on *Model Characterization*. The goal of this exercise was to identify the time varying piece wise constant functions for *c*, *q*, *β* and *ϕ*.*pI* that result in best visual fit of the cumulative infections on the log scale. The focus was on reproducing the predominant features of data sets on the log scale such as, piecewise linear slopes, steepening of slopes, plateauing effects and changing y-intercepts with constant slope. We did not use any empirical learning methods for discovery of these parameters as we were keen on developing intuition regarding the relation between trend patterns and their underlying processes. Once the cumulative data of infections was well fit, no further changes in parameters were undertaken except for tuning *δ* in order to bring the curve of cumulative deaths within the band defined by the curves *D_q_* and *D_q_* + *D*. This band was necessitated because deaths reported are sometimes from one or both buckets. Ever since the Indian government has mandated the testing of all Severe Acute Respiratory infections for COVID-19, undetected infections have been confirmed positive for COVID-19 from time to time in ICUs and post mortem. A fixed reporting delay delay of 8 days was used for calculating recoveries. This is observed in empirical explorations of data from Indian States

### Model characterization

In order to explain the performance of the model and capture the dynamics we administered several scenarios to the model. Each of the scenarios are explained below. In each of the scenarios, we set *a* = 1 implying that epidemic is in stage 2. Thus efficiency of intervention is given by 1 − (1 − *c*)^2^. All the discussion relating to c, efficiency of intervention (1 − (1 − *c*)^2^) or inefficiency of intervention ((1 − *c*)^2^) in succeeding discussions apply equally to *q* (if transmission is community transmission), 1 − (1 − *c*)[*a*.(1 − *c*) + (1 − *a*)(1 − *q*)] and (1 − *c*)[*a*.(1 − *c*) + (1 − *a*)(1 − *q*)]

#### Effect of varying *β* and *c* with fixed *R_t_*

It is well known that as *R_t_* increases, the infections peak earlier and have higher peaks. But from the equation 1 we can see that same *R_t_* can be achieved by varying combinations of *β* and intervention inefficiencies. To understand this effect, we fixed *R_t_* at 3 and varied numerators of transmission rate (*β*) were set successively at values between 6 and 8.4. *c* was varied to obtain required *R_t_* as per Eqn. 1 in the *Model parameters* section. From Eqn. 1, we can observe that at constant *R_t_*, increase in 0 will have to be accompanied by a decrease in inefficiency (1 − *c*)^2^ and a consequent increase in efficiency (1 − (1 − *c*)^2^) and detection *c*. Latency of infection is set at 7 days, efficiency of random testing and self-presentation (q) is set at 0, influx of population (*ϕ* * *pI*) is set at 0. Initial infected cluster is set at 1 for a state with a population of one million and it is assumed that epidemic is at contact transmission stage (Stage-2).

#### Varying R_t_ by variations of β and interventions

We vary *R_t_* from 5/3 through 7/3 in steps of 0.5/3. We achieve these reproduction numbers, by varying either *β* or *c*.

To understand the effect of varying β when intervention (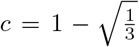) is held constant, we simulated the model with the above-mentioned parameters but with *β* values of 5, 5.5, 6, 6.5 and 7. The other parameters were held at the same value as the previous section.

Next, we study the effect of varying intervention (*c*) when *β* = 6/9 is held constant, we simulated the model with the above-mentioned parameters but with *c* values as: 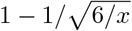 where *x* ∊ {5/3, 5.5/3, 6/3, 6.5/3, 7/3} thereby achieving required *R_t_*. The other parameters were held at the same value as the section on *Effect of R_t_*.

#### Effect of influx of infected cohort

To understand the effect of influx of infected persons we simulated an influx of infected over a small window of 3, 9 and 18 days. We set the influx rate to 38 people per day (*ϕ* = 75; *pI* = 0.5) during the influx window. The other parameters were held at the same value as the section on *Effect of R_t_*.

#### Transitory effects of varying β and c over short time windows

*β* and interventions sometimes change transitionally over short periods of time. Lock-downs, focused campaigns of contact tracing or testing are examples.

To study the effect of varying *β* and *c* over short periods, the scenarios as follows. Starting with a *β* = 6/9 and 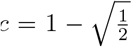 we apply an (i) increase in β over a 21 day window to 8/9 and a (ii) pulsed decrease in β for 21 days to 4/9. All other parameters were held at the same value as the previous section. Thus starting from a base *R_t_* = 3, the *R_t_* was stepped up and down respectively to 4 and 2 respectively.

To study similar effects induced by c, starting from the same baseline values, we apply (i) an increased 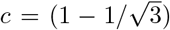 over a 21 day window (*R_t_* = 2) and a (ii) pulsed decrease in *c* for 21 days to (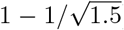) (*R_t_* = 4). The other parameters were held at the same value as the section on *Effect of R_t_*.

#### Effects of intervention on influx of infections

We coupled an influx of infections with varying levels of intervention and with varying delays to study how they affect the overall evolution of the epidemic. We started with a *β* = 5/9 and a *c* = 0.2 and applied an influx at the rate of 20 per day for 5 days. Interventions with different efficiencies were applied on the first day of influx using *c* = 0.4, 0.6, 0.8 successively. Additionally, we also applied an intervention *c* = 0.8 starting 5 days after the end of the influx to assess the effect of delay in intervention.

## Results

### Model characterization

#### Effect of interventions with fixed Rt

Fig. 3 presents the model responses for *R_t_*=3 respectively. Increased *β* requires proportional increase in *c* to keep *R_t_* constant (Refer Eqn. 1). At fixed *R_t_*, while the time course of the epidemic remains the same, the total numbers of infected increase with increasing beta. But due to commensurate increase in *c*, the excess infections are sucked into the pool of detected and quarantined infections. Since only the undetected infections influence transmission, *β_eff_*, *R_t_* and time course of epidemic remain same, while the peak of reported infections are higher as can be seen in Fig.3(a).

**Figure 3.**
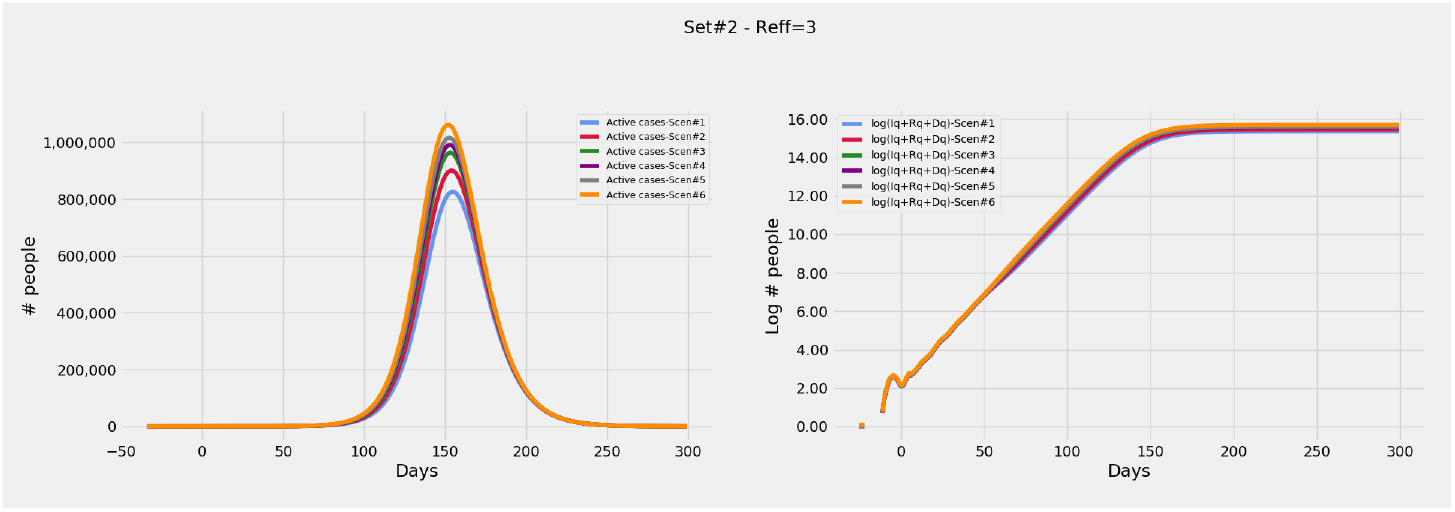
Effect of *β* and *c* at constant *R* = 3. *R_eff_* is set to 3. Plots showing (a) Evolution of active cases of infected persons, (b) Cumulative number of cases over a 300 day epoch on log scale.

#### Varying R_t_ by variations of β and interventions

Fig. 4 a and b presents the model responses to varying interventions and constant *β*, while Fig. 4c and d depict model responses to varying *β* at constant interventions. All simulations start with the same parameters and the variations are executed midway on day 50. It may be seen from Fig. 4 that increase in *R_t_* effected by increase in *β* or decrease of *c* results in steepening of the slope in Fig. 4b and d and earlier peaks in Fig. 4a and c. Similarly decreasing *R_t_* decreases the slope and later peaks. However at the same *R_t_*, the peak values of infections are proportional to *β*. This is evident from the Fig. 4c where grey and indigo curves have taller peaks, but red and blue curves have shorter peaks compared to corresponding peaks in 4a. Note that the correspondence is between the pairs of curves in gray and blue, red and indigo.

**Figure 4.**
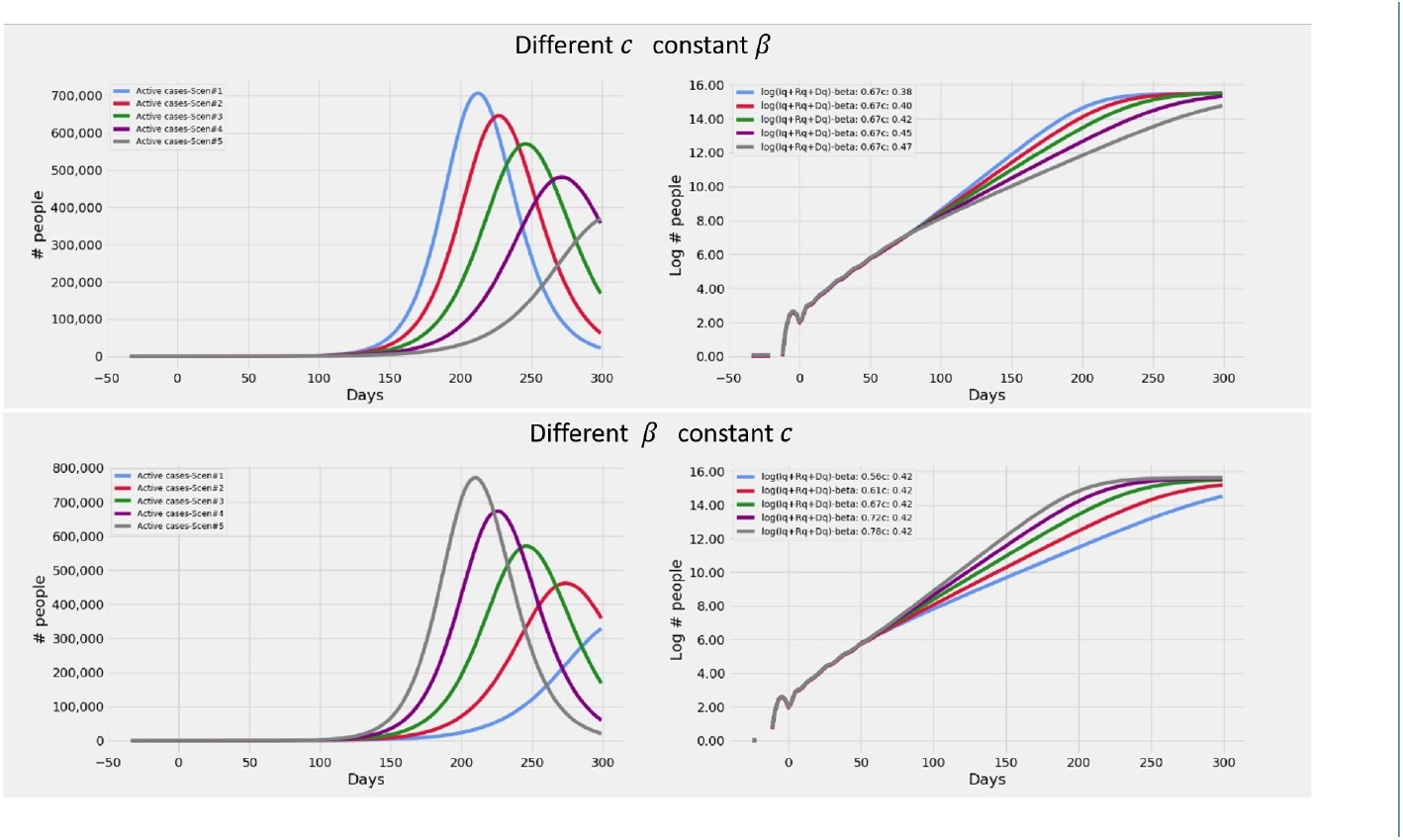
Effect of varying *β* with a constant intervention. Plots showing **(a)** Evolution of active cases of infected persons at various *c* and constant *β* **(b)** Cumulative number of cases over a 300 day epoch on log scale at various *c* and constant *β***(c)** Evolution of active cases of infected persons at various *β* and constant *c* **(d)** Cumulative number of cases over a 300 day epoch on log scale at various *β* and constant *c*. All simulations start with the same values of *β* and *c*. The variations of *β* and *c* are applied on day 50 (See Methods for values of parameters used)

#### Effect of influx of infected cohort

Fig. 5 describes the effects of an influx of infected individuals into the population. It can be seen that this influx does not change the slope of the trajectory. Rather, the trajectory maintains its slope, but now has an increased y intercept. Influxes over windows of 3, 9 and 18 days have progressively larger y intercepts as seen in Fig. 5b and the peaks occur earlier in time as seen in Fig. 5a. However the change in y intercepts are not uniform. Another significant effect is that the excess infections due to this influx are constant on the log scale. Thus with time, the cost of this influx become progressively larger in absolute terms.

**Figure 5.**
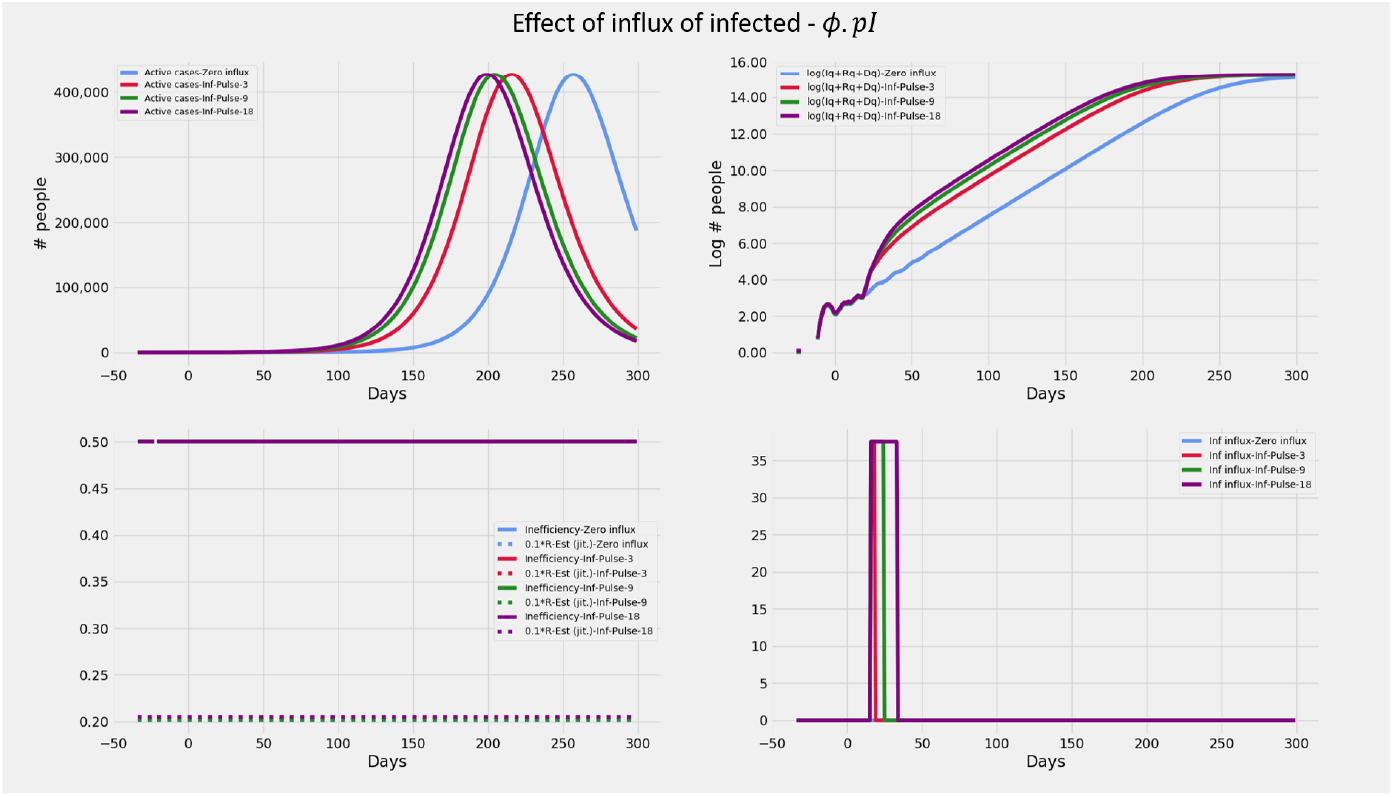
Effect of pulsed influx infected persons. Plots showing **(a)** Evolution of active cases of infected persons, **(b)** Cumulative number of cases over a 300 day epoch on log scale. (See Methods for details)

#### Transitory effects of varying β and c over short time windows

The effect of variations in *β* and *c* on the slopes of cumulative infections on log scale are similar to that mentioned in section on *Effect of varying R_t_* during the short window of variation. However at the end of the window, the trajectory is restored back to its original slope and now proceeds parallel to the original trajectory. Thus the net effect of change in *R_t_* in a short window results in a change in y intercept akin to that of an influx of infected cohort. This effect is clearly seen in Fig. 6b and d where the decrease or increase of y-intercept is evident. It may also be noted that the effect of β sets in almost instantaneously, while that of c shows up after a short lag. It may be explained as the lag required for the reduction in undetected infections to show up as a decrease in effective transmission.

**Figure 6.**
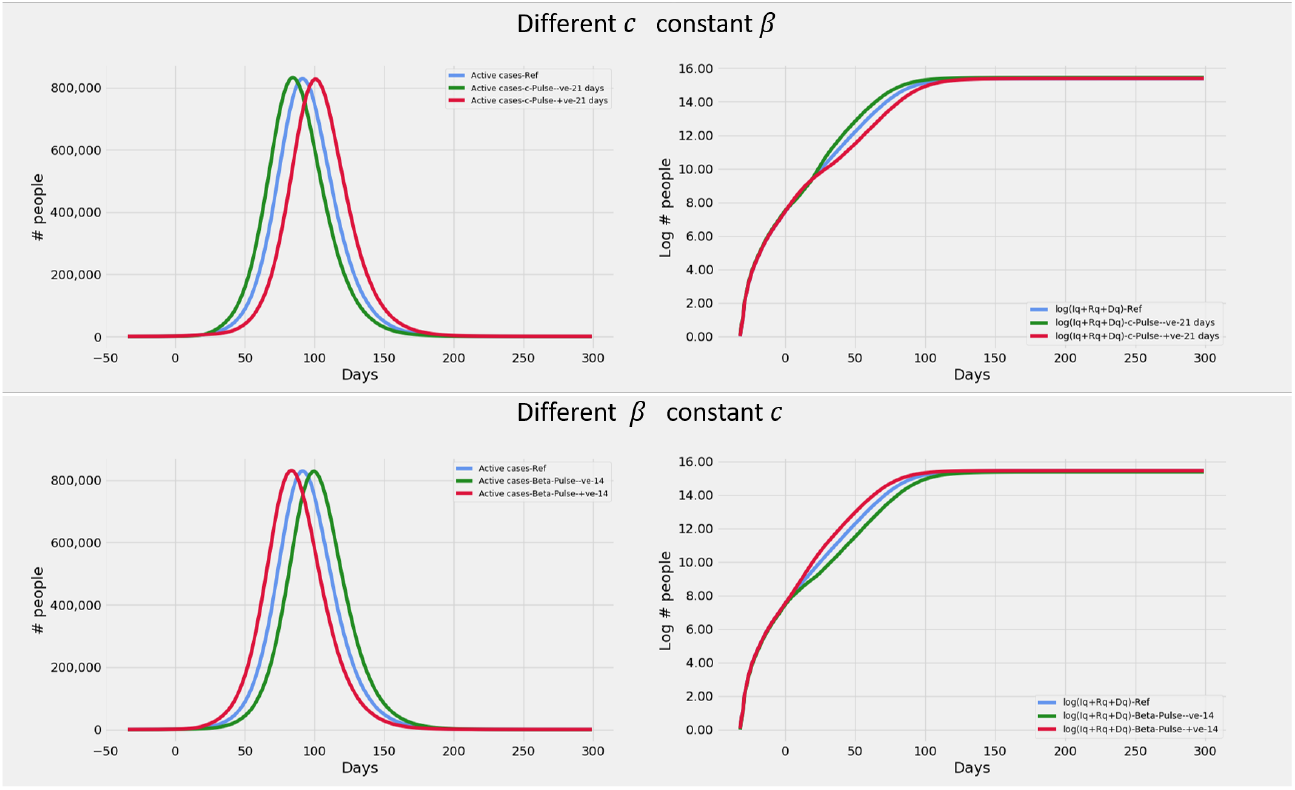
Transitory effects of varying *β* and *c* over short time windows. Plots showing **(a)** Evolution of active cases of infected persons at various c and constant *β* **(b)** Cumulative number of cases over a 300 day epoch on log scale at various c and constant *β* **(c)** Evolution of active cases of infected persons at various *β* and constant *c* **(d)** Cumulative number of cases over a 300 day epoch on log scale at various *β* and constant *c*. All simulations start with the same values of *β* and *c*. The variations of *β* and *c* are applied on day zero (See Methods for values of parameters used)

#### Effects of intervention on influx of infections

The effects of contact tracing as a response to influx of infected is elucidated in Fig. 7. As already seen in section on *Effect of infected cohort*, an influx accompanied by no response in the form of change in contact tracing efficiency results in a trajectory that runs parallel to the original trajectory with increased y intercept. However if as a response to the influx, the level of contact tracing is increased concurrent with the first day of influx, it leads to a peculiar 2 phase change in trajectory. There is a steep increase in slope almost instantaneously on increase of *c*, followed by a significant reduction in slope with respect to the original slope. With increasing levels of contact tracing *c*, the sharp increase in the initial phase is even steeper, and so is the flattening that follows. The initial steep rise can be attributed to large numbers of infections being detected and quarantined due to increased contact-tracing efforts. This results in a drastic reduction in the slope soon after, as quarantined infected cannot spread the disease. This effect leads to a characteristic plateauing effect. When the intervention is delayed, the plateau effect seen is identical in nature but shifted in time. Hence the resultant flattening is achieved at a higher level of infections.

### Heuristics for reconstructing the time varying interventions in real data

The salient features seen in real data of cumulative infections on the log scale have been reproduced in the previous sections. These features are:

- Piece wise linearity
- Changes in slope in each piece wise segment
- Temporary deviations due to transitory change in slope resulting in parallel trajectories with altered y intercept
- Plateau effect: Sudden rise followed by a flattening

**Figure 7.**
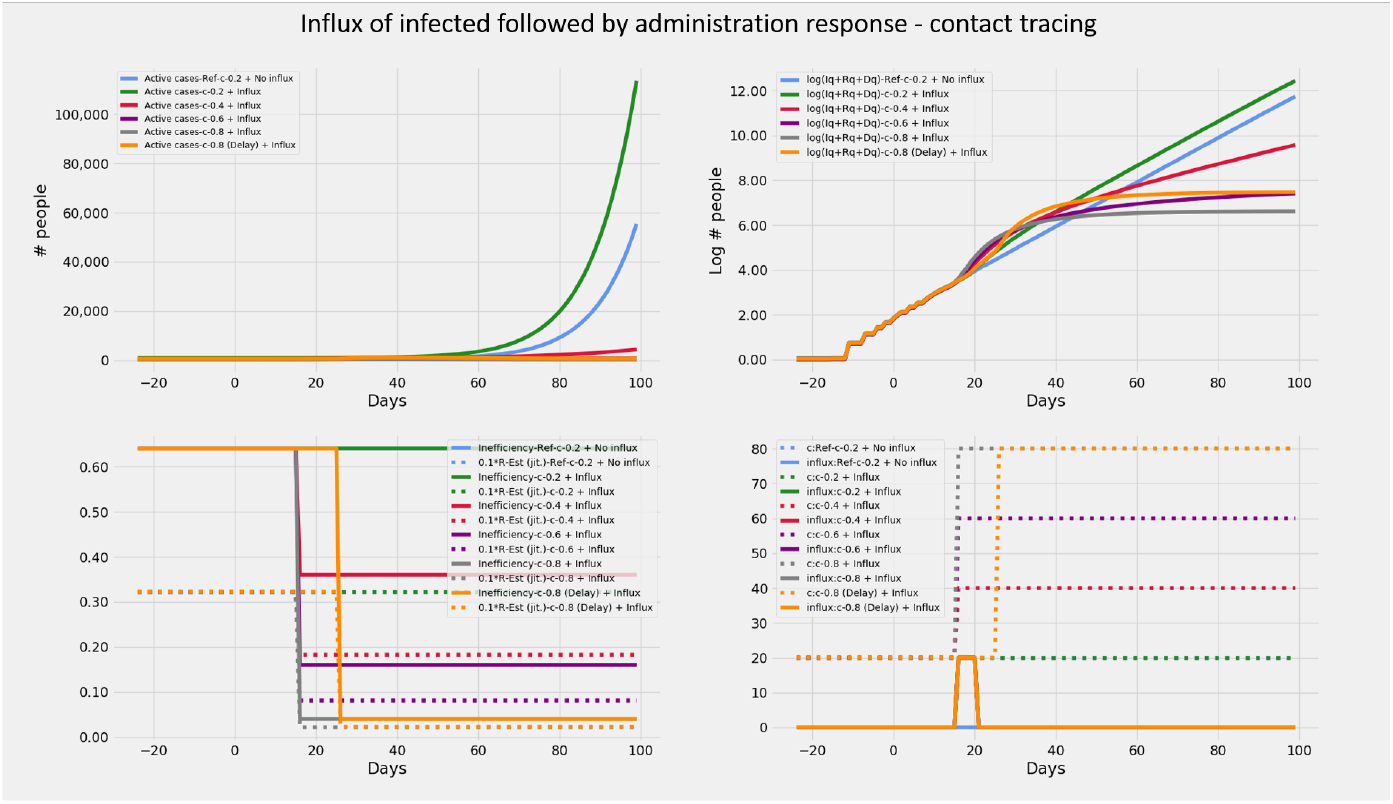
Effect of influx of infected followed by contact tracing exercises with various efficacies. Plots showing **(a)** Evolution of active cases of infected persons and its response in the form of varying intensities and timing of intervention **(b)** Cumulative number of cases over a 100 day epoch on log scale **(c) R_t_** and *interventionin efficiency* over the duration of the epidemic **(d)** Time course of the influx of infected given by *ϕ*(*t*)*.pI*(*t*) and *c*. (See Methods for values of parameters used). Notice the green, red, indigo and gray lines exhibit progressively stronger plateau effect. The effect of delayed intervention in orange runs parallel to the gray line. The cost of the delay is the difference in the y intercept between the orange and gray lines on the log scale.

Based on the results encountered so far, we propose the following set of heuristics to reconstruct using our model, the observed trajectory of cumulative infections in real data:

1. The first piece of the piece wise linear curve is reconstructed by using a suitable combination of *β* and *c*. This indirectly fixes the underlying *R_t_*. As seen in section on *Effect of interventions with fixed R_t_* although there are multiple combinations of *β* and *c* can recreate the *R_t_*, each of these combinations has a unique trajectory of active infections.
2. For each subsequent piece of the piece wise linear curve, modify *β* and / or *c* to match the slope of the piece
3. For each parallel shift of trajectory introduce an influx of infected. While it is possible to account for this using a transitory change in *β* or *c* as well, we assume that transitory and self restoring changes in *β* or *c* over short periods are not very likely except in the case of lock downs. Thus we use population influx as the first choice tool to account for this feature
4. For steep rise followed by a significant flattening in the span of a short interval, we use an influx with increased heightening of contact tracing levels *c*.

### Modelling epidemic data from Indian states

Based on the heuristics arrived at in section on *Heuristics for reconstructing interventions* we recreate the curves of cumulative infection, recovery and death on the log scale for twelve Indian states. In order to recreate, we set *c*, *q*, *β* and *ϕ.pI* as a piecewise constant time varying function based on the heuristics described in the previous section. This activity is performed only on the data up to -10 days. No further changes in parameters are allowed after this day. But the model generates outputs upto day zero. The model output for the days -10 to zero are *model predictions*.

The piece wise functions are set in order to recreate the cumulative infections only. No explicit attempt is made to recreate the trends of recoveries and deaths except the following.

1. The recoveries are shifted right by a fixed lag of 8 days for all states.
2. A single value of mortality (deaths per infection) is chosen for each state such that the curve of real data(dark gray) is bounded by the two model generated cumulative death curves (by counting deaths from only detected infections, and by counting deaths from both detected and undetected infections).

A few general comments may be in order on the agreement between model predictions and real data.

1. The model predictions for the last 10 days agree very well with the true data
2. For a recovery to be declared, at least 2 successive negative tests are required. It is likely that states are being extremely conservative in declaring recoveries and want to err on the side of safety. This could explain the lag of about 8 days which is required to fit the real data with the model generated output.
3. The real recoveries seem to follow the model generated curves, but with a staircase effect. This effect could possibly be due to a procedural issue where test reports are processed in batches on a slightly lower priority(which recoveries are compared to critical cases).

The full details of the piece wise constant functions used for each state in reconstruction and the reasoning applied to generate the same can be found in the supplementary materials. Supplementary materials also include an evaluation of the predictions based on the same model for an epoch of 20 days upto 7^th^ May 2020. It can now be seen that as expected there are significant deviations in prediction for 5 states. This is expected as small underlying changes in the intervening period widen the gap between real data and prediction. But it is also significant that for more than half the states considered, predictions held well over a 20 day epoch. However in general we believe that this model must be evaluated weekly to recognise and register the underlying dynamic changes in the epidemic progress. This exercise also provides inputs on efficacy of interventions and new influxes or new clusters that have emerged over the week.

## Discussion

This work presents a modified SEIRD model with ability to represent population influx and interventions including contact tracing and random testing. This model was used to show that some of the commonly seen features of cumulative infections in real data can be explained by piece wise constant changes in interventions and population influx. We also show that the data of cumulative infections from twelve Indian states between mid March and mid April 2020 can be generated from the model by applying interventions according to a set of heuristic rules. The model with interventions so designed was allowed to generate prediction data for ten more days with no further modifications. This predicted trend matched well with the data for the twelve states. Matching the infections curve ensured that the recoveries and deaths too matched well with no additional effort.

**Figure 8.**
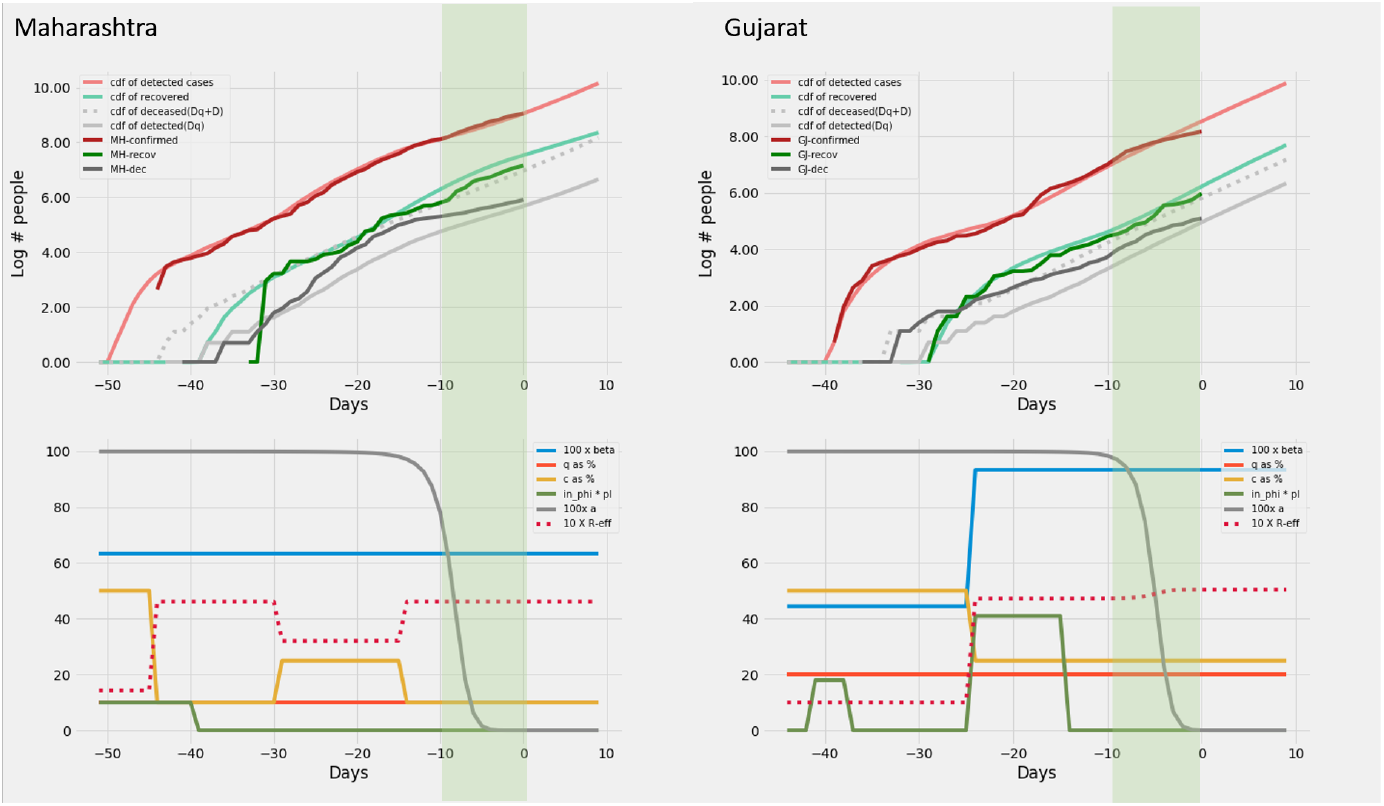
Maharashtra and Gujarat: Recreation. Plots on the top row show the real and model generated curves, while the bottom row plots the time varying piece wise functions *c*, *q*, *β* and *ϕ.pI* used to recreate the same. The piece wise functions were created in accordance with the heuristic algorithm described in section on *Heuristics for reconstructing interventions*. On the top row, dark colours indicate the real data from the states, while the lighter shades are model generated. The red shades are cumulative infections, green shades are recoveries and grays are deaths. The dotted light gray line indicates the curve output from the model if deaths from both the undetected and undetected infections are counted. The light gray solid line counts only the deaths from detected infections in the model. The band in green highlights the prediction epoch. Day zero corresponds to April 27^th^ 2020.

**Figure 9.**
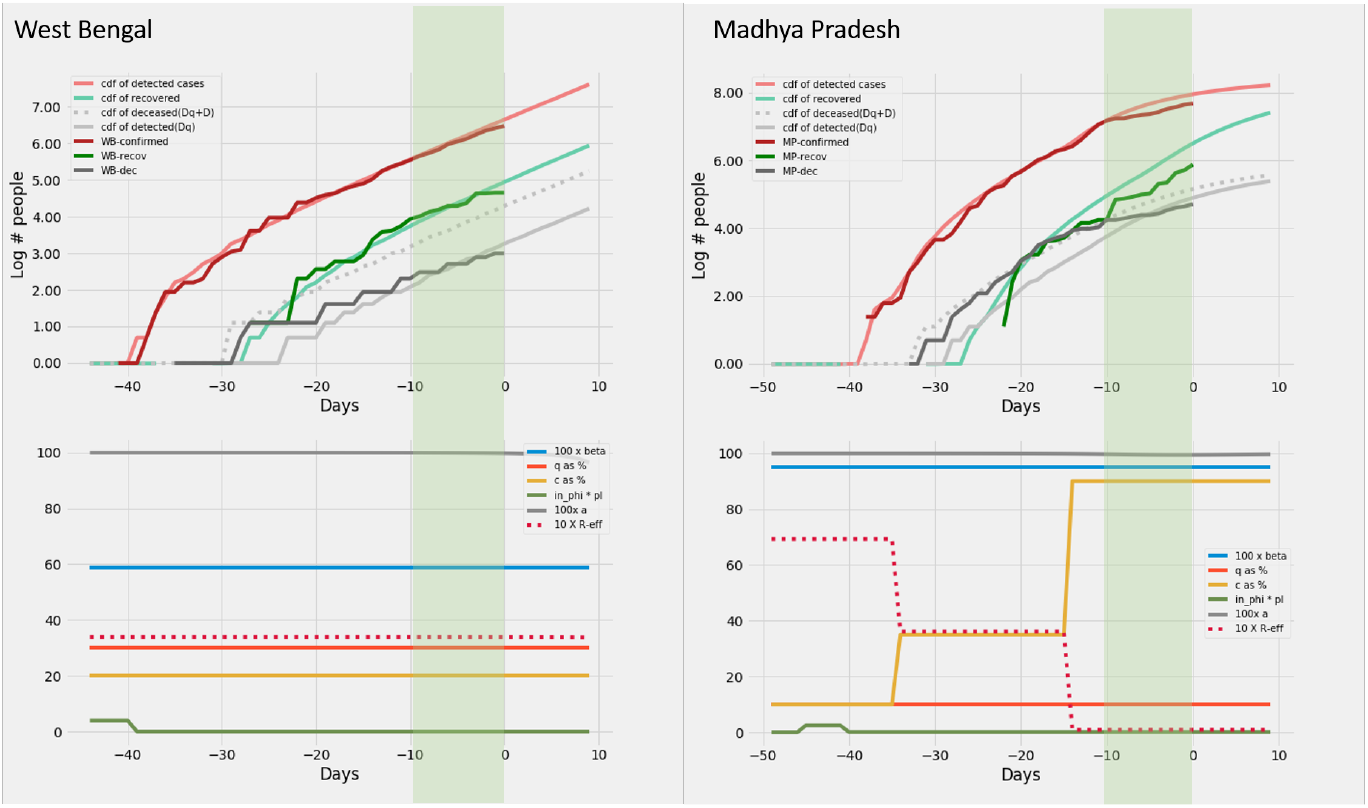
West Bengal and Madhya Pradesh: Recreation. Plots on the top row show the real and model generated curves, while the bottom row plots the time varying piece wise functions *c*, *q*, *β* and *ϕ*.pI used to recreate the same. The piece wise functions were created in accordance with the heuristic algorithm described in section *Heuristics for reconstructing interventions*. On the top row, dark colours indicate the real data from the states, while the lighter shades are model generated. The red shades are cumulative infections, green shades are recoveries and grays are deaths. The dotted light gray line indicates the curve output from the model if deaths from both the undetected and undetected infections are counted. The light gray solid line counts only the deaths from detected infections in the model. The band in green highlights the prediction epoch. Day zero corresponds to April 27^th^ 2020.

**Figure 10.**
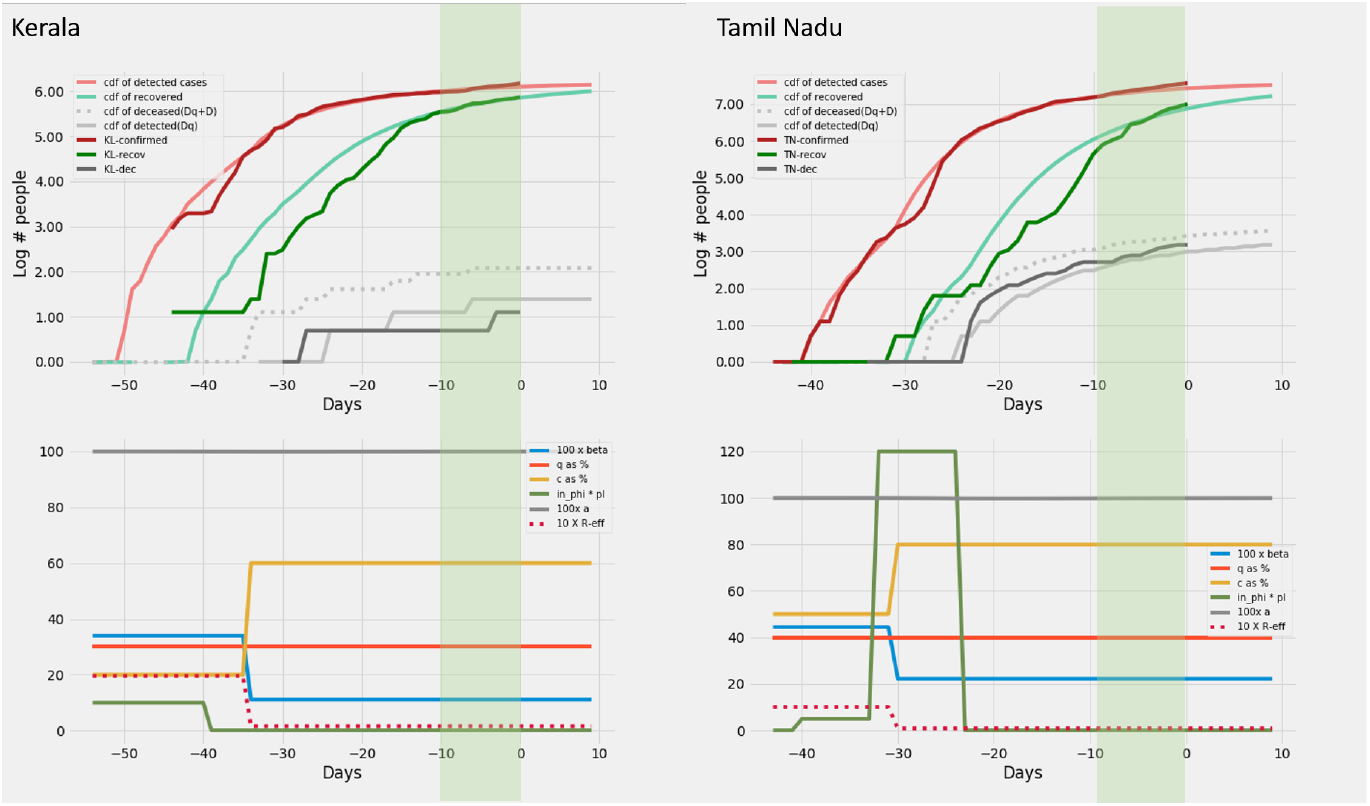
Kerala and Tamil Nadu: Recreation. Plots on the top row show the real and model generated curves, while the bottom row plots the time varying piece wise functions *c*, *q*, *β* and *ϕ*.*pI* used to recreate the same. The piece wise functions were created in accordance with the heuristic algorithm described in section on *Heuristics for reconstructing interventions*. On the top row, dark colours indicate the real data from the states, while the lighter shades are model generated. The red shades are cumulative infections, green shades are recoveries and grays are deaths. The dotted light gray line indicates the curve output from the model if deaths from both the undetected and undetected infections are counted. The light gray solid line counts only the deaths from detected infections in the model. The band in green highlights the prediction epoch. Day zero corresponds to April 27^th^ 2020.

**Figure 11.**
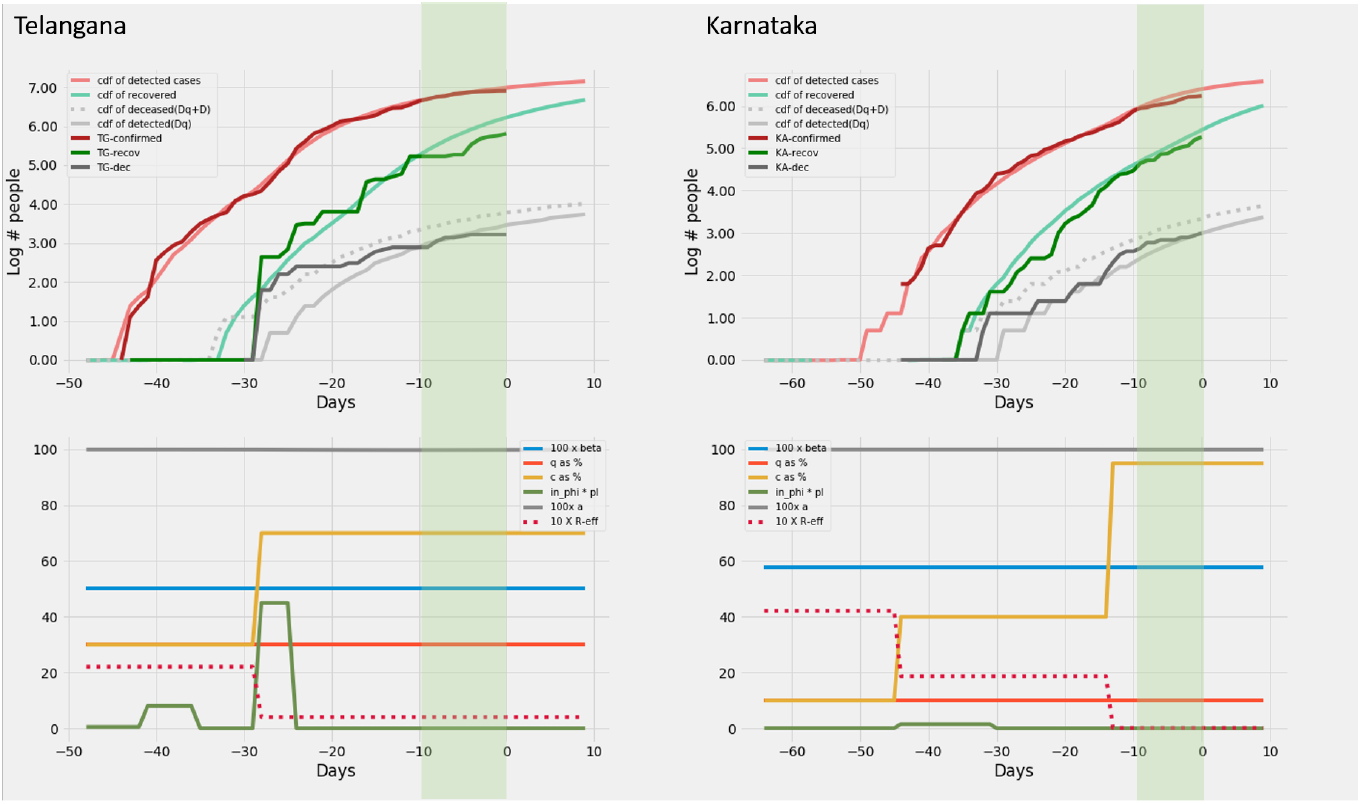
Telangana and Karnataka: Recreation. Plots on the top row show the real and model generated curves, while the bottom row plots the time varying piece wise functions *c*, *q*, *β* and *ϕ*.*pI* used to recreate the same. The piece wise functions were created in accordance with the heuristic algorithm described in section on *Heuristics for reconstructing interventions*. On the top row, dark colours indicate the real data from the states, while the lighter shades are model generated. The red shades are cumulative infections, green shades are recoveries and grays are deaths. The dotted light gray line indicates the curve output from the model if deaths from both the undetected and undetected infections are counted. The light gray solid line counts only the deaths from detected infections in the model. The band in green highlights the prediction epoch. Day zero corresponds to April 27*^th^* 2020.

**Figure 12.**
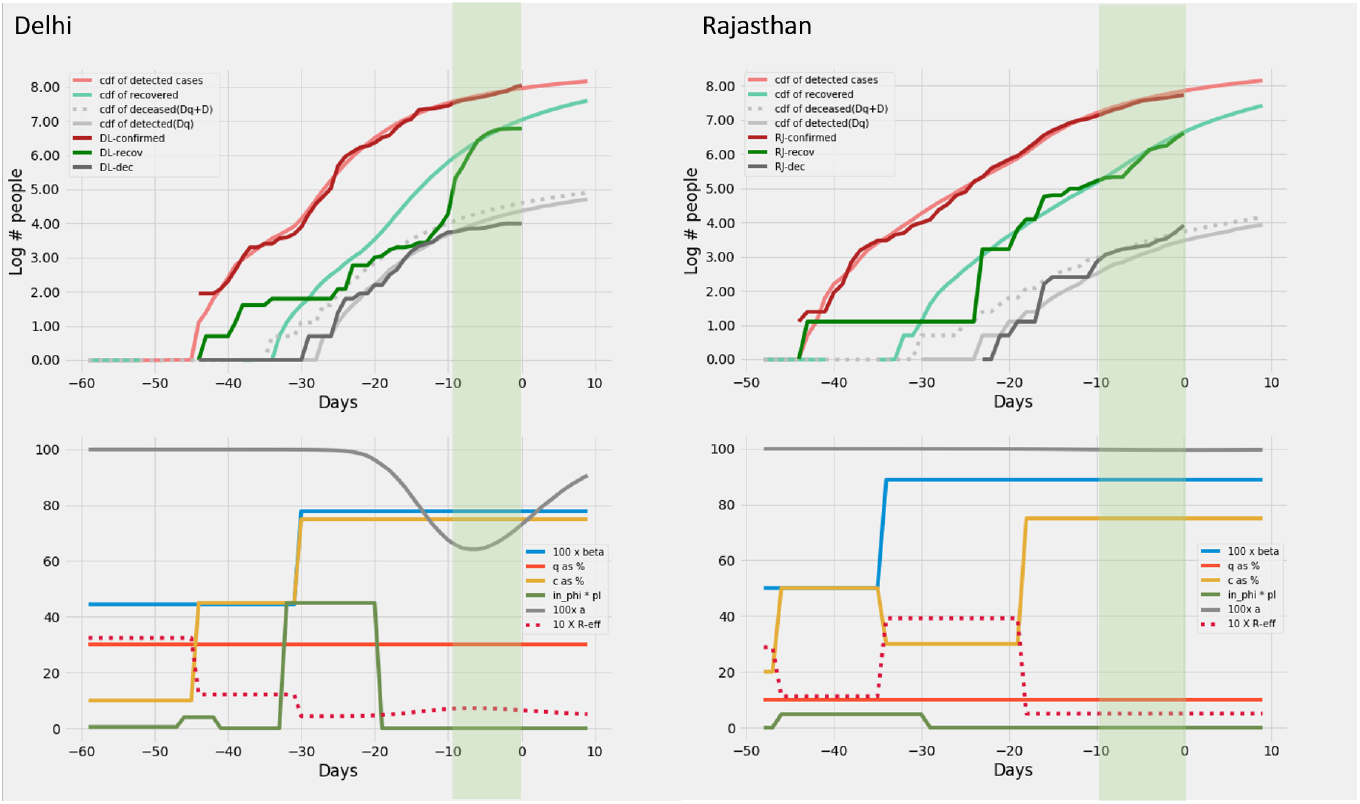
Delhi and Rajasthan: Recreation. Plots on the top row show the real and model generated curves, while the bottom row plots the time varying piece wise functions *c*, *q*, *β* and *ϕ.pI* used to recreate the same. The piece wise functions were created in accordance with the heuristic algorithm described in section *Heuristics for reconstructing interventions*. On the top row, dark colours indicate the real data from the states, while the lighter shades are model generated. The red shades are cumulative infections, green shades are recoveries and grays are deaths. The dotted light gray line indicates the curve output from the model if deaths from both the undetected and undetected infections are counted. The light gray solid line counts only the deaths from detected infections in the model. The band in green highlights the prediction epoch. Day zero corresponds to April 27*^th^* 2020.

**Figure 13.**
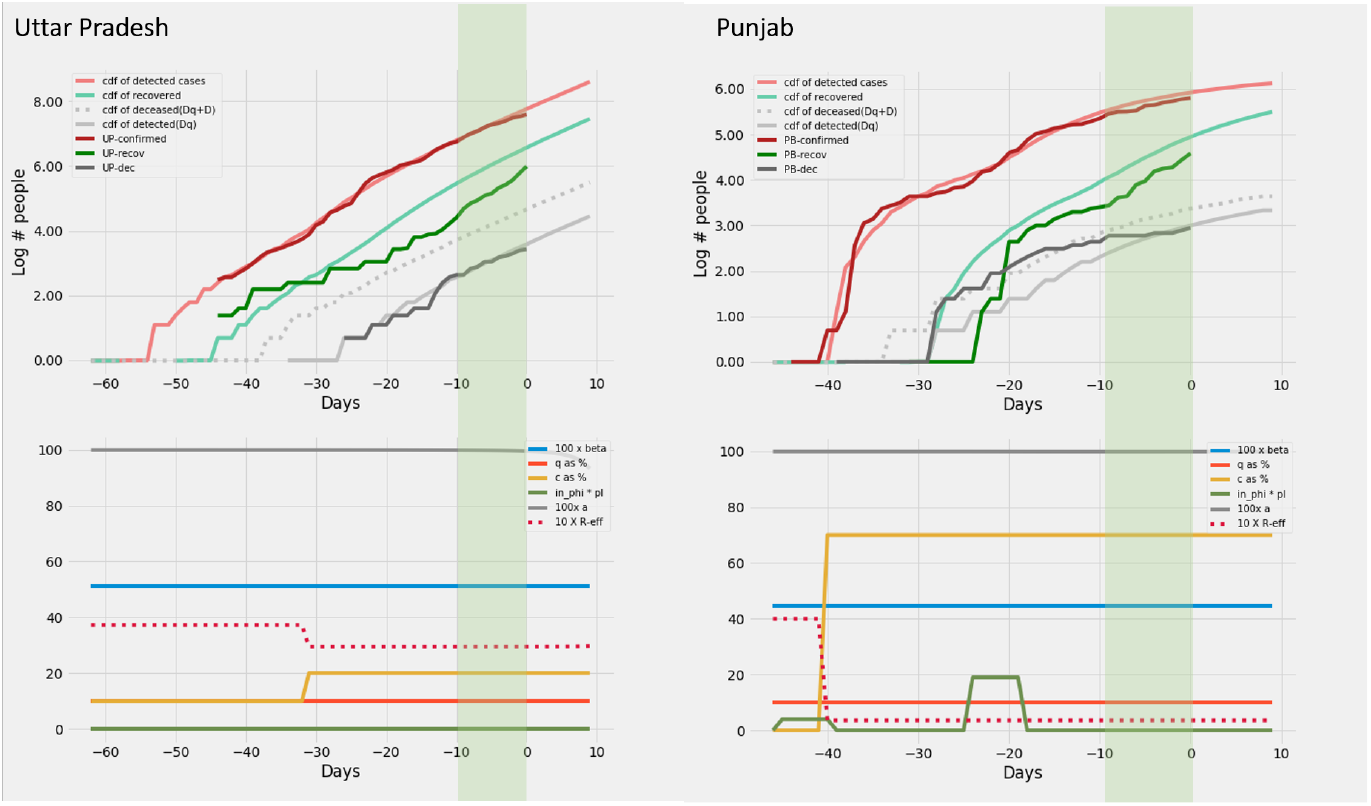
Uttar Pradesh and Punjab: Recreation. Plots on the top row show the real and model generated curves, while the bottom row plots the time varying piece wise functions *c*, *q*, *β* and *ϕ*.*pI* used to recreate the same. The piece wise functions were created in accordance with the heuristic algorithm described in section on *Heuristics for reconstructing interventions*. On the top row, dark colours indicate the real data from the states, while the lighter shades are model generated. The red shades are cumulative infections, green shades are recoveries and grays are deaths. The dotted light gray line indicates the curve output from the model if deaths from both the undetected and undetected infections are counted. The light gray solid line counts only the deaths from detected infections in the model. The band in green highlights the prediction epoch. Day zero corresponds to April 27*^th^* 2020.

We thus provide a mechanistic model for short term prediction of COVID-19 epidemic data. Compared to empirical measures [14, 19, 17] this method recreates data using parameters that are intuitive and directly related to operation on the field. While similar to other mechanistic models [3, 4, 16], it differs in its design focus on separating observable and non observable compartments. Connecting elementary piece wise changes in interventions to features of real data like slopes and plateaus are a singular contribution of this work. While the piece wise constant form of interventions have been proposed [14], instead of creating interventions functions based on known events and identifying their contributions to data fit, we compose the piece wise interventions directly based on intuitive connections between data features and elementary piece wise functions of interventions.

The model discussed in this manuscript signifies efforts, not to merely model the evolution of the epidemic but also captures the effects of various interventions by the administration. It presents a method by which administration may get an objective estimate of several aspects of an ongoing epidemic such as the efficacy of contact tracing apparatus, possible influx of infected, extent of social distancing or transmission. The parameters c and q as fraction of infected detected are generic enough for a wide variety of cases. While administrations may use a wide variety of methods to characterize their testing or tracing strategies, their eventual success lies in tracing or detecting all infected, which is what is captured by c and q. Thus insights from our model provides an independent and objective feedback on how effective the efforts have been. This model also provides an estimate of R*t* which is highly sought after and provides insights on whether the observed R*t* results from interventions or transmission. The population influx mechanisms provide a way for administrations to estimate the effect of influx in the past and plan for impending arrivals and calibrate their responses.

A few notes may be in order in interpreting the results from the methods proposed in this work. The insights coming out of our models must be interpreted in the context of the specific clusters contributing to the epidemic. For instance increased or unchanged transmission does not imply that general population flouted lock down rules. It must instead be interpreted specifically as applicable to the specific clusters of spread. *β* is interpreted as the product of contact rate and probability of transmission. Thus large captive households or even hospitals can keep the contact rates high without any flouting of norms. Again, influx of infected in its general sense represents a seeding of infection. Thus an influx of population indicated by our results could as well imply the emergence of a new infection cluster.

The primary intention behind building the model is to monitor localized response using dashboards. This can be possible with better data availability at the local level (district in the case of India). Our model design principles are generic enough to be applied to model extensions incorporating hospitalized or critical cases. Since the model captures the active cases as a function of response of the administrative response, this can be a valuable tool to plan and allocate resources and deploy the right response, where necessary. This model can further be extended to estimate the effort required for achieving a level of contact tracing *c* or testing *q*. This can be of vital importance for the administration in planning their strategies and estimating requirements of health workers, law enforcement agencies for contact tracing. We currently manually fit the parameters to evolution of active cases in each of the 10 states using heuristics. Going forward, the manually obtained piece wise interventions may be treated as seed for empirical discovery of piece wise functions. Further incorporating confidence intervals for our current predictions will be an important goal of future work.

## Data Availability

All epidemiological data used was obtained from public repositories of www.covid19india.org

## Competing interests

The authors declare that they have no competing interests.

## Notes

### Competing Interest Statement

The authors have declared no competing interest.

### Clinical Trial

Using publicly available data

### Funding Statement

No funding was received for this study.

## References

1. Kermack, W.O., McKendrick A G.: A contribution to the mathematical theory of epidemics (1927). doi:10.1002/mma.5067

2. Ronald, R.: Constructive Epidemiology. The British Medical Journal 1(3562), 673 (1929)

3. Lipsitch, M., Cohen, T., Cooper, B., Robins, J.M., Ma, S., James, L., Gopalakrishna, G., Chew, S.K., Tan, C.C., Samore, M.H., Fisman, D., Murray, M.: Transmission dynamics and control of severe acute respiratory syndrome. Science 300(5627), 1966–1970 (2003). doi:10.1126/science.1086616

4. Kissler, S.M., Tedijanto, C., Goldstein, E., Grad, Y.H., Lipsitch, M.: Projecting the transmission dynamics of SARS-CoV-2 through the post-pandemic period. medRxiv (March), 2020–030420031112 (2020). doi:10.1101/2020.03.04.20031112

5. Hufnagel, L., Brockmann, D., Geisel, T.: Forecast and control of epidemics in a globalized world. Proceedings of the National Academy of Sciences of the United States of America 101(42), 15124–15129 (2004). doi:10.1073/pnas.0308344101

6. Lemon, D.M., Apostolakis, G.E.: Massachusetts Institute of Technology Engineering Systems Division A Methodology for the Identification of. Technology (2004)

7. Diekmann, O., Heesterbeek, J.A.P., Metz, J.A.J.: On the definition and the computation of the basic reproduction ratio R0 in models for infectious diseases in heterogeneous populations. Journal of Mathematical Biology 28(4), 365–382 (1990). doi:10.1007/BF00178324

8. Heesterbeek, J.A.P.: A brief history of R0 and a recipe for its calculation. Acta biotheoretica 50(3), 189–204 (2002)

9. Heffernan, J.M., Smith, R.J., Wahl, L.M.: Perspectives on the basic reproductive ratio. Journal of the Royal Society Interface 2(4), 281–293 (2005). doi:10.1098/rsif.2005.0042

10. Diekmann, O., Heesterbeek, J.A.P., Roberts, M.G.: The construction of next-generation matrices for compartmental epidemic models. Journal of the Royal Society Interface 7(47), 873–885 (2010). doi:10.1098/rsif.2009.0386

11. Mizumoto, K., Chowell, G.: Transmission potential of the novel coronavirus (COVID-19) onboard the diamond Princess Cruises Ship, 2020. Infectious Disease Modelling 5(February), 264–270 (2020). doi:10.1016/j.idm.2020.02.003

12. Wallinga, J., Lipsitch, M.: How generation intervals shape the relationship between growth rates and reproductive numbers. Proceedings of the Royal Society B: Biological Sciences 274(1609), 599–604 (2007). doi:10.1098/rspb.2006.3754

13. Grassly, N.C., Fraser, C.: Mathematical models of infectious disease transmission. Nature Reviews Microbiology 6(6), 477–487 (2008). doi:10.1038/nrmicro1845

14. Flaxman, S., Mishra, S., Gandy, A., Al, E.: Imperial College London. Faculty of Medicine. COVID-19 reports. Report 13: Estimating the number of infections and the impact of non-pharmaceutical interventions on COVID-19 in 11 European countries. Imperial College COVID-19 Response Team (March), 1–35 (2020)

15. Wallinga, J., Teunis, P.: Different epidemic curves for severe acute respiratory syndrome reveal similar impacts of control measures. American Journal of Epidemiology 160(6), 509–516 (2004). doi:10.1093/aje/kwh255

16. Singh, R., Adhikari, R.: Age–structured impact of social distancing on the COVID-19 epidemic in India, 1-9 (2020). 2003.12055

17. Kucharski, A.J., Russell, T.W., Diamond, C., Liu, Y., Edmunds, J., Funk, S., Eggo, R.M., Sun, F., Jit, M., Munday, J.D., Davies, N., Gimma, A., van Zandvoort, K., Gibbs, H., Hellewell, J., Jarvis, C.I., Clifford, S., Quilty, B.J., Bosse, N.I., Abbott, S., Klepac, P., Flasche, S.: Early dynamics of transmission and control of COVID-19: a mathematical modelling study. The Lancet Infectious Diseases 3099(20), 1–7 (2020). doi:10.1016/S1473-3099(20)30144-4

18. Snehal Shekatkar, Bhalchandra Pujari, Mihir Arjunwadkar, Dhiraj Kumar Hazra, Pinaki Chaudhuri, Sitabhra Sinha, Gautam I Menon, Anupama Sharma, V.G.: INDSCI-SIM A state-level epidemiological model for India (2020). https://indscicov.in/indscisim

19. Prakash, M.K., Kaushal, S., Bhattacharya, S., Chandran, A., Kumar, A., Ansumali, S.: A minimal and adaptive prediction strategy for critical resource planning in a pandemic. medRxiv, 2020–040820057414 (2020). doi:10.1101/2020.04.08.20057414

